# The application of Raman spectroscopy and chemometric methods for analysis healthy blood and blood with BRCA mutation

**DOI:** 10.1101/2022.02.21.22271291

**Authors:** M. Kopec, B. Romanowska-Pietrasiak, H. Abramczyk

## Abstract

Presented study included human blood from heathy patients and from patients with BRCA mutation. Raman spectroscopy can be used for BRCA mutation detection and bioanalytical characterization of pathologically changed samples. The aim of this study is to evaluate the Raman biomarkers to distinguish blood samples from healthy patients and from patient with BRCA mutation by Raman spectroscopy. We have proved that Raman spectroscopy is a powerful technique to distinguish between healthy blood and blood with BRCA mutation and to characterize biochemical composition of samples. Partial Least Squares Discriminant Analysis yielded effective and comparable samples classification based on vibrational features. The sensitivity and specificity obtained from PLS-DA have been over 86.5%. The obtained results confirm clinical potential of Raman spectroscopy in oncological diagnostics.

**Graphical Abstract:** 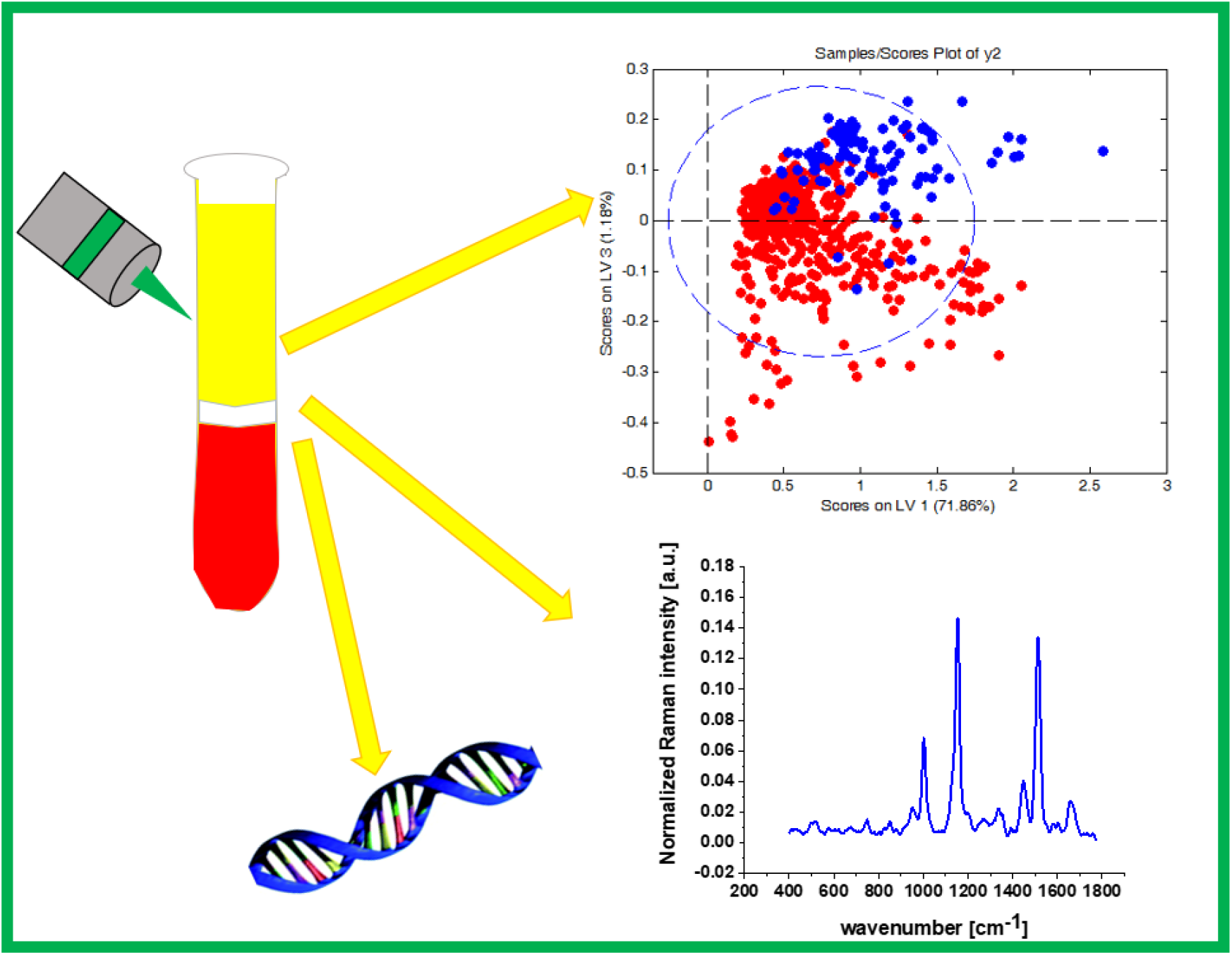

## Introduction

Blood is important fluid that circulate through the body and has a lot of functions that are fundamental for surviving [1] The first function of blood is transportation with supplying oxygen to all cells and tissues. Moreover, blood supplies essential substances to cells, such as sugars or hormones and removes waste products from cells. [2] Second function is regulation such as body right temperature or concentration each of the components of the blood [3]. Third function of blood is protecting the body from diseases and infections. [2] [3] [4] Blood transfers a lot of molecular information and reflects real health based on metabolism. [5] In blood there is a lot of biomarkers that can be useful tool for characterizing pathology processes including cancer. [6] [7] [8] [9] [10] Blood marker analysis may help identify early stages of pathological processes that can lead to future serious diseases. [11]

Breast cancer is one of the most common cancer in population. Moreover it is the most common cause of cancer death. [12] Breast cancer genes (BRCA) belong to the group which produce proteins that help repair DNA. [13] [14] People who inherit pathogenic modifications of BRCA have increased risk of cancers, especially breast cancer. [15] [16] There are studies demonstrating that tumor grade correlates with the level of BRCA proteins. [17]

The potential of BRCA proteins expression in breast cancer has been studied by numerous groups. [18] [19] [20] [21] [22] [23] [6] E. Gross et al. have used chromatography to study BRCA mutation. [24] They have compared results for BRCA mutation obtained by direct sequencing method with results obtained by high performance liquid chromatography (DHPLC). They reported that DHPLC is fast, sensitive and powerful method for genetic analysis. [24] [25] J. Roth et al. have used Ultra HPLC with Tandem Mass Spectrometric Detection (uHPLC-MS/MS) to analyse activity Olaparib in human plasma. [26]

Recently next generation sequencing (NGS) technologies have been widely used for BRCA mutation analysis. [27] [28] E. Szczerba et al. have studied BRCA mutation in the tumor tissue using NGS. [29] They used bioinformatic NGS software programs to analyse BRCA. [29] [21] NGS procedures can become an invaluable tool for clinical diagnosis in the near future.

The BRCA gene test is a standard method used for people who have an inherited mutation based on family history. The results of this method aren’t always clear. So it is justified to find new methods for detection and treatment of early-stage disease. This hope may be provided Raman spectroscopy. To the best of our knowledge it is only few paper described using Raman spectroscopy to study BRCA mutation. L.R. Allain et al. for the first time have used SERS to monitor DNA hybridization of a fragment of the BRCA1 breast cancer susceptibility gene on modified silver surfaces. [30] They proved that SERS technique for gene expression studies is a significant step forward in enabling biological quantification. [30] Another group which used SERS to analyze BRCA mutation was M. Culha et al. [31] Raman spectroscopy is a tool that provides information about biochemical composition of organelles in single cells. Raman spectroscopy allows to identifying cancer biomarkers which can discriminate between normal state and cancer pathology.

In this paper, we present that Raman spectroscopy combined with statistical methods of artificial intelligence made possible to discriminate between healthy patients and patient with BRCA mutation by testing human blood plasma

In this paper, we focus on the analysis of Raman spectra of blood plasma from patients without BRCA mutations (control) and patients with BRCA mutations using the optimal multivariate methods to differentiate between samples. Raman spectroscopy gives a new promise to provide a perfect tool for BRCA mutation diagnosis.

## Materials and methods

### Sample preparation

The blood samples were obtained from the Voivodeship Multi-Specialist Center for Oncology and Traumatology in Lodz. The procedure did not affects the course of the treatment of the patients. Written informed consent was obtained from all patients, or if subjects are under 18, from a parent and/or legal guardian. All experiments were conducted in accordance with relevant guidelines and regulations of the Bioethical Committee at the Medical University of Lodz, Poland (RNN/17/20/KE). The experimental protocols were approved by Bioethical Committee at the Medical University of Lodz, Poland (RNN/17/20/KE). All the experiments were carried out in accordance with Good Clinical Practice and with the ethical principles of the Declaration of Helsinki. Spectroscopic analysis did not affect the scope of course and type of undertaken hospital treatment.

Patients with BRCA mutation were diagnosed and treated at the Voivodeship Multi-Specialist Center for Oncology and Traumatology in. For all experiments we used human fresh blood. Blood samples were collected in Ethylene Diamine Tetra Acetic Acid (EDTA) vials and next centrifuged at 3500 rpm for 5 min in 18°C to yield the plasma samples. A 10 µl drop of each plasma blood was placed on clean CaF2 windows (Crystran).

### Raman spectroscopy

The Raman measurements were recorded using an alpha 300 RSA+ (WITec, Germany) combined with confocal microscope coupled via the fiber of 50 μm core diameter with a spectrometer UHTS (Ultra High Throughput Spectrometer) and a CCD Camera (Andor Newton DU970N-UVB-353) operating in standard mode with 1600 × 200 pixels at −60 °C with full vertical binning. Standard calibration procedure was performed every day before measurement with the use of silicon plate (520.7□cm^−1^). Raman data were pre-processed were performed using the WITec Project Plus Software. Each Raman spectra were processed to remove cosmic rays. The corrected Raman spectra were smoothed by a Savitzky and Golay procedure and baseline subtraction. All experiment were performed using laser beam (SHG of the Nd:YAG laser (532 nm)) and 40x dry objective (Nikon, objective type CFI Plan Fluor CELWD DIC-M, numerical aperture (NA) of 0.60 and a 3.6–2.8 mm working distance). Raman spectra for plasma blood samples were recorded using integration time 1 and 10 number of accumulations. All experiments were performed using laser with a power was 10 mW at the sample position.

The background subtraction and the normalization (model: divided by norm (divide the spectrum by the dataset norm)) were performed by using Origin software. The normalization model: divided by norm was performed according to the formula:

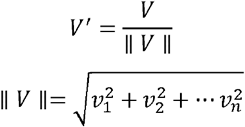

where:

*v*_*n*_ is the n^th^ V values. [32]

The normalization was performed for spectral region (400-1700 cm^-1^)

### Molecular methods for the detection of BRCA1 and BRCA2 germline mutations

Molecular diagnosis of genetic disorders is defined as searching for and revealing defects in DNA and/or RNA samples. Detection of mutations in the BRCA1 and BRCA2 genes involves several sequencing procedures. The methods for identification of the disease-causing mutations can be classified as methods for detection of known mutations and those unknown [33] [34] [35].

#### First procedure

The first step in performing DNA extraction is sampling (blood, saliva or tissue specimens, etc.). Peripheral blood (most frequently used to obtain DNA for human genetic studies) can be obtained using tubes treated with EDTA or Sodium Citrate to prevent clotting. Then, the human genomic DNA is isolated from fresh or frozen blood or cells and identified, isolated and subsequently purificated [36,37]

#### Second procedure

In families carrying a pathogenic variant in BRCA1 and BRCA2 genes usually Sanger sequencing method has been used for detecting them. It is the gold-standard DNA sequencing technique. This method is based on the selective incorporation of chain-terminating dideoxynucleotides by DNA polymerase during in vitro DNA replication. Traditional Sanger sequencing is restricted to the discovery of small defects (substitutions, insertions, deletions). This technology plays a major role in verification of PCR results [38,39]

#### Third procedure

The third stage of testing is screening for the point known mutations. Polish studies showed that the BRCA1 founder effect exist with a predominant presence of variants such as - c.5266dupC, c.181T>G, c.4035delA, c.68_69delAG, c.3700_3704delGTAAA, but the mutations spectrum is more heterogeneous. The main technique dedicated to the detection of known mutations in BRCA1 and BRCA2 is the polymerase chain reaction (PCR) and its variations, for example Real Time PCR. This method is characterized by high sensitivity and specificity [40–42]

#### Fourth procedure

In clinical cases of high probability of unknown mutations identification the next-generation sequencing (NGS) technique is used. This is a new technology used for DNA and RNA sequencing and detection of different types of genes mutations such as known, unknown, founder or recurrent [43–45].

#### Fifth procedure

The exons deletions or amplifications related cases of hereditary breast cancer might not be detected by sequence analysis. In that case another molecular diagnostic technique should be used. Multiplex Ligation-dependent Probe Amplification (MLPA) offers the possibility to detect large genomic rearrangements [46–49].

### Statistical analysis

We have analyzed samples with BRCA mutation from 85 patients and samples from control patients without BRCA mutations from 15 patients. In total, samples from 100 patients have been analyzed. In detail, for samples with BRCA mutations we used typically 460 Raman single spectra, for samples from healthy patients we used 104 Raman spectra.

Statistical chemometric analysis was performed by using Mathlab and PLS_Toolbox Version 4.0. Partial Least Squares Discriminant Analysis (PLSDA) was used for building predictive classification models, to validate the classification models and to calculate sensitivity and specificity. A ROC curve analysis has also been performed. More details about chemometric methods was described in our previous papers. [50,51]

## Results and discussion

In this section we will present the results for human blood plasma from patients without BRCA mutations (control) and patients with BRCA mutations. Figure 1 A and B presents average normalized Raman spectra of human plasma with standard deviation (dark shadows) based on 104 single spectra (control) and 460 spectra of patients with BRCA mutations. Figure 1C presents difference Raman spectrum (human blood plasma with BRCA mutation - control).

**Fig 1.**
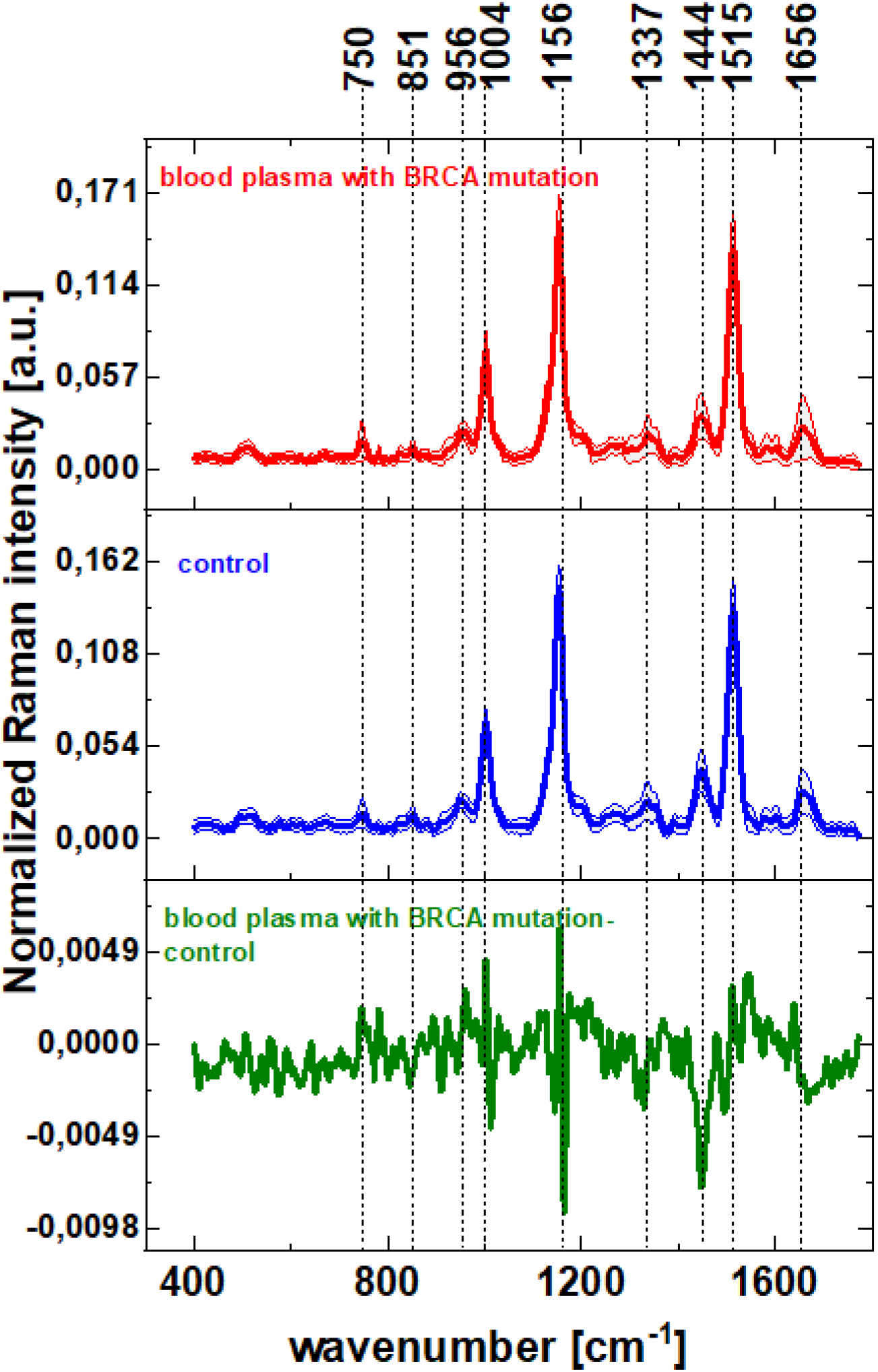
The normalized average Raman spectra typical for: (A) plasma from healthy patients (control) with SD (blue line), (B) plasma from patients with BRCA mutation with SD (red line), (C) the difference spectrum (blood plasma with BRCA – control) (green line)

One can see from Fig 1 A and B that Raman spectra of the human blood plasma can be characterized by vibrational peaks at: 750, 851, 956, 1004, 1156, 1337, 1444, 1515 and 1656 cm^-1^. In Table 1 we present the tentative assignments of Raman peaks observed in human blood plasma.

**Table 1.** Tentative assignments of Raman peaks [52] [53] [54] [55] [56]

| Wavenumber [$\text{cm}^{-1}$] | Tentative assignments |
| --- | --- |
| 750 | Tryptophan, cytochrome c, hemoglobine |
| 851 | tyrosine |
| 956 | hydroxyproline/Collagen backbone |
| 1004 | phenylalanine |
| 1156 | carotenoids |
| 1337 | cytochrome c |
| 1444 | fatty acids, triglycerides $\text{CH}_2$ or $\text{CH}_3$ |
| 1515 | carotenoids |
| 1656 | Amid I, lipids |

One can see from the difference spectrum at Fig. 1C that peak at 1004 cm^-1^ assigned to phenylalanine and 750 cm^-1^ corresponding to cytochrome c/hemoglobine are positive, what confirms the higher amount of cytochrome c in blood plasma with BRCA mutation. The positive difference is also observed at 1656 cm^-1^. This peak is assigned to amid I/lipids which is stronger in the Raman spectra for blood plasma with BRCA mutation (positive correlation in difference spectrum).

Comparing the peaks at 1444 cm^-1^ assigned to fatty acids/triglycerides CH_2_ or CH_3_ one can see that this band is stronger in Raman spectrum for healthy blood plasma (negative correlation in difference spectrum). The same tendency is observed for peaks at 1156 cm^-1^ and 1515 cm-1 assigned to carotenoids. This observation is consistent with our previous papers for cancer pathology. [51] [57] [58]

In order to visualize chemical similarities and differences between human blood plasma for patients without BRCA mutations and blood plasma samples for patients with BRCA mutation we have evaluated the predictive validity and robustness of Raman spectroscopy by using multivariate statistical methods for data interpretation. We have performed statistical analysis for 564 single spectra. To simplify the task of analyzing a large number of samples and multidimensional Raman vectors (intensities vs. wavenumbers) were subjected to dimension reduction by means of Partial Least Squares Discriminant Analysis (PLSDA). The PLSDA analysis was applied to the 460 Raman spectra of 85 human blood samples with BRCA mutation and from the 104 Raman spectra of 15 human blood samples without BRCA.

By plotting the principal component scores, similarities between the samples can be revealed. Figure 2 shows the PLSDA score plot for the Raman spectra of the human blood plasma samples with BRCA mutations and for the samples without BRCA mutations.

**Fig 2.**
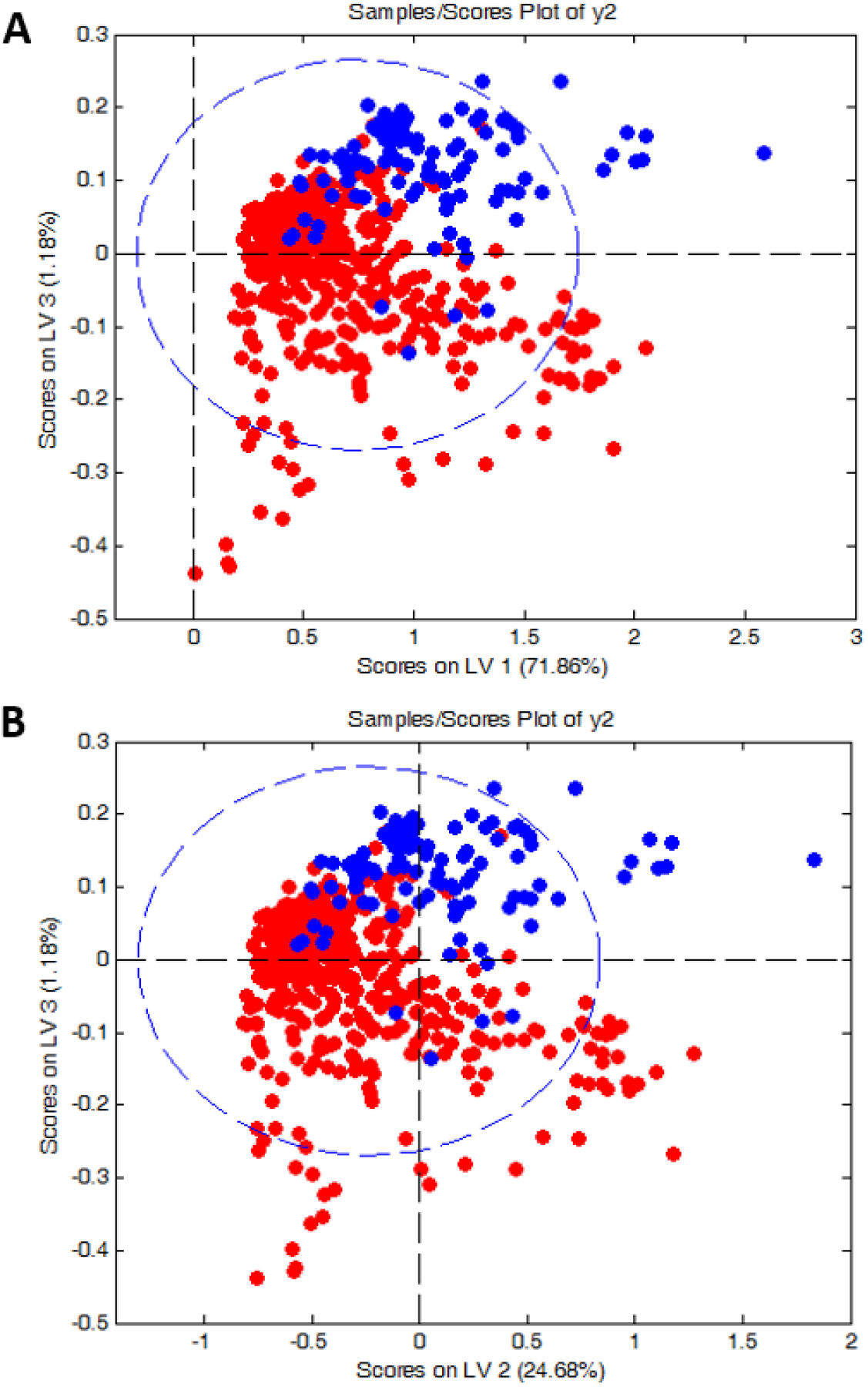
The scores plots obtained from PLSDA for the Raman spectra of blood plasma without BRCA mutations (blue circle) and for Raman spectra of blood plasma with BRCA mutations (red circle)

One can see from Fig. 2 that evident differences between human blood plasma with BRCA and for human blood plasma without BRCA mutations are revealed. The differences are visible by grouping the results into two separate clusters. Raman spectra for human blood plasma without BRCA mutations (blue circle) are grouped in the right upper area of the plot while the samples from human blood plasma with BRCA mutations (red circle) are grouped in the lower area of the plot. The plot presented on Fig. 2 confirms the differences between samples for human blood plasma without BRCA mutations and for human blood plasma with BRCA mutations.

To understand the molecular information contained in the LV1, LV2 and LV3 we used the loading plots presented in Fig. 3 that reveal the most important characteristic features in the Raman spectra.

**Fig 3.**
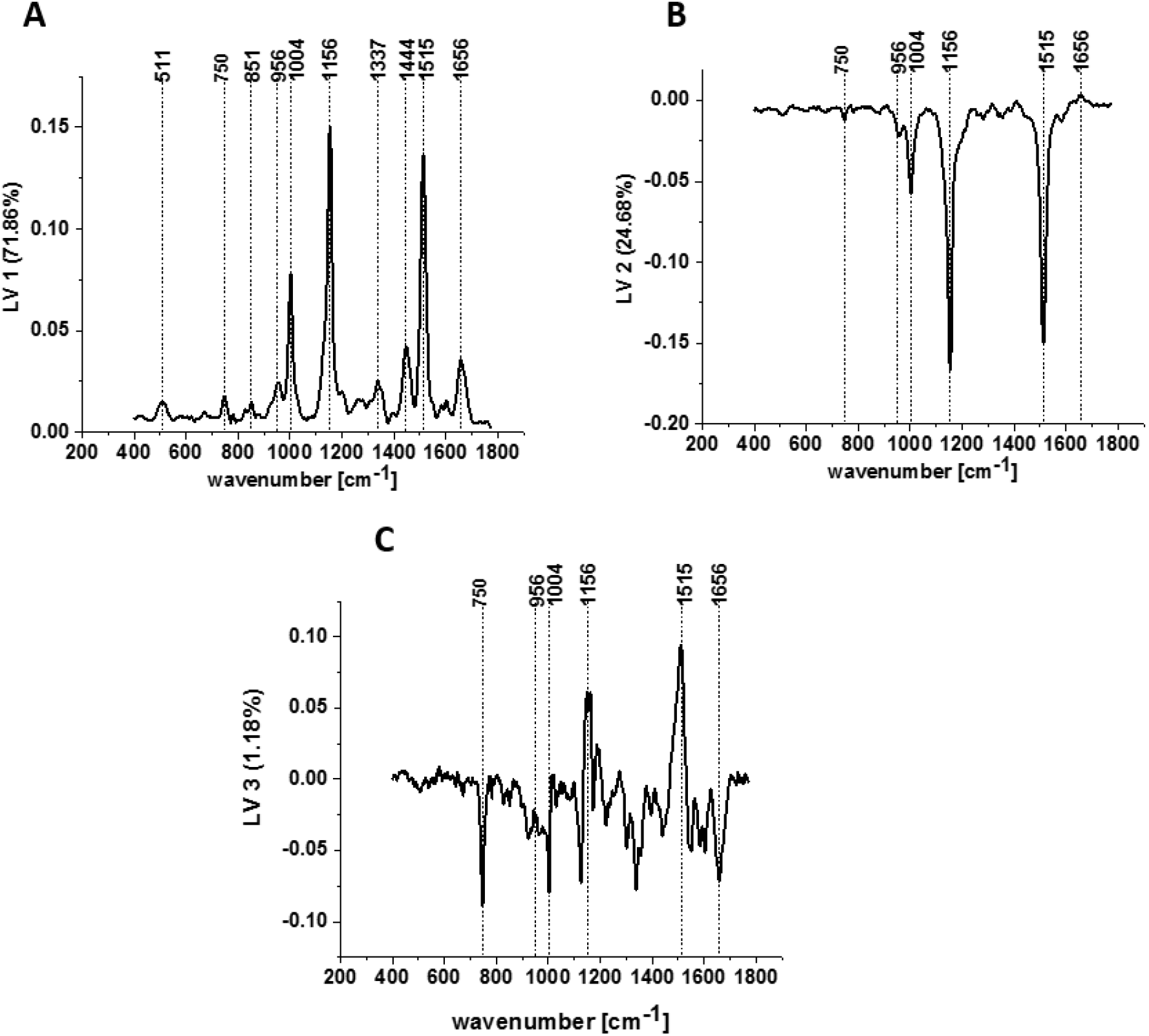
PLSDA loading plot for LV1 (A), LV2 (B), LV3 (C) for the Raman spectra of human blood plasma without BRCA mutations and for Raman spectra of human blood plasma with BRCA mutation.

Figure 3 shows the loading plots of LV1, LV2 and LV3 versus wavenumbers obtained from PLSDA methods for two classes of spectra typical for human blood plasma without BRCA mutations and for human blood plasma with BRCA mutation. One can see that the loading plots show the most pronounced changes around the characteristic Raman peaks of carotenoids and proteins.

Once can see from Fig. 3 that the first LV has the contribution of 71.86%, LV2 the contribution of 24.68% and LV3 the contribution of 1.18% to variance, respectively. LV1 and LV2 give the dominant account for the maximum variance in the data.

The first latent variable (LV1) is presented in Fig. 3 A. The most characteristic maxima in the loading plot are at 511 cm^-1^, 750 cm^-1^, 851 cm^-1^, 956 cm^-1^, 1004 cm^-1^, 1156 cm^-1^, 1337 cm^-1^, 1444 cm^-1^, 1515 cm^-1^, and 1656 cm^-1^. The second latent variable LV2 (Fig. 3B), reaches its minima at 750 cm^-1^, 956 cm^-1^, 1004 cm^-1^, 1156 cm^-1^, 1515 cm^-1^ and 1656 cm^-1^. The third latent variable is presented in Fig. 3C and reaches its maxima at 1156 cm^-1^and 1515 cm^-1^ and minima at 750 cm^-1,^ 1004 cm^-1,^ 1656 cm^-1^.

Table 2 presents the results of calculation of the sensitivity and specificity obtained from PLS-DA method.

**Table 2.** The values of sensitivity and specificity for calibration and cross validation procedure from PLS-DA analysis.

|  | Healthy blood plasma | Blood plasma with BRCA mutation |
| --- | --- | --- |
| Sensitivity (calibration) | 0.913 | 0.885 |
| Specificity (calibration) | 0.885 | 0.913 |
| Sensitivity (cross validation) | 0.900 | 0.865 |
| Specificity (cross validation) | 0.865 | 0.900 |

Figure 4 presents ROC curves (Receiver Operating Characteristic) curves for blood plasma sample without BRCA mutations and for blood plasma sample with BRCA mutations. Figure 4 confirms high potential of Raman spectroscopy to differentiate between patients without BRCA mutations and with BRCA mutations.

**Fig 4.**
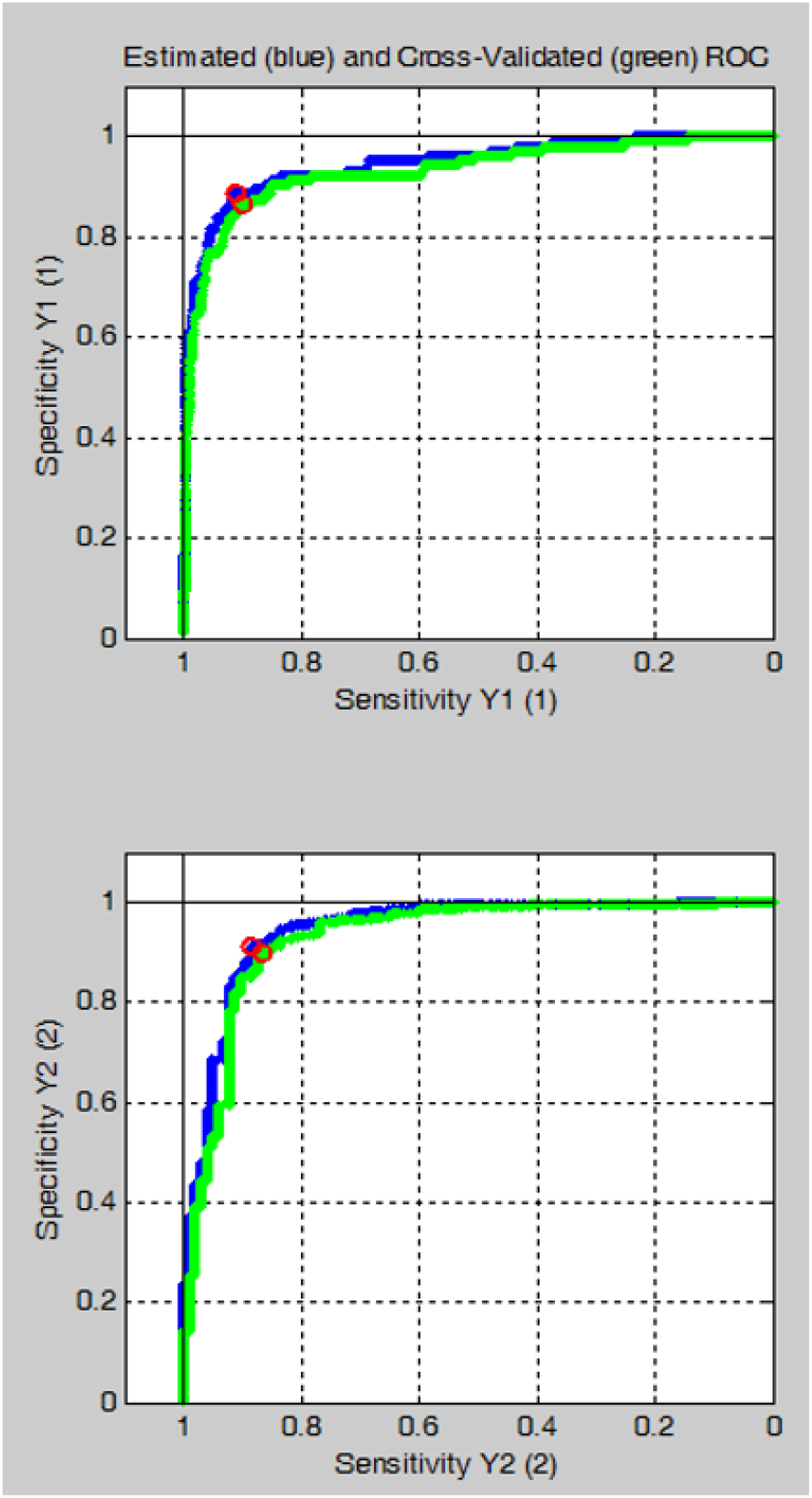
ROC curves obtained from the PLS-DA analysis for two classes of Raman spectra assigned to blood plasma without BRCA mutations and blood plasma with BRCA mutation.

## Conclusions

In this paper we used Raman spectroscopy to monitor BRCA mutations in human blood plasma. Raman spectroscopy and PLSDA have been shown to be excellent tool for the diagnosis of breast BRCA mutations. Raman spectroscopy and chemometric method have been successfully applied to characterize and differentiate human blood plasma without BRCA mutations and human blood plasma with BRCA mutation. The results presented in this paper suggest that Raman biomarkers in the near future can provide additional insight into the biology of human blood plasma. The differentiation by Raman spectroscopy between blood plasma without BRCA mutations and blood plasma with BRCA mutations is important because the high specificity and sensitivity can lead to genomic breast diagnosis. Our results can help implement Raman spectroscopy as a tool for blood analysis to investigate BRCA mutations.

## Data Availability

All data produced in the present study are available upon reasonable request to the authors

## Acknowledgement

This work was supported by the National Science Centre of Poland (Narodowe Centrum Nauki, UMO-2019/33/B/ST4/01961).

## CRediT authorship contribution statement

Conceptualization: H.A., M.K.; Funding acquisition: H.A., Investigation: M.K., B.R-P., Methodology: M.K., B.R-P.; Writing-original draft: M.K., H.A.; Manuscript editing: M.K., H.A. All authors have read and agreed to the published version of the manuscript.

